# Engaging Racialized Newcomers in Chronic Disease Prevention and Management: A Scoping Review of Health Promotion Interventions

**DOI:** 10.1101/2025.08.29.25334665

**Authors:** Marisa L. Kfrerer, Dawn P Gill, Claire Styffe, Paul Severiano Aspinall, Ariana Tang, Justin Ovadia, Callie Hirsh, Robert J Petrella

**Affiliations:** Health & Rehabilitation Sciences, Western University, London ON, N6A 3K7, Canada; Centre for Studies in Family Medicine, Department of Family Medicine, Schulich School of Medicine & Dentistry, Western University, London, ON, N6G 2M1, Canada; Department of Family Practice, Faculty of Medicine, University of British Columbia, Vancouver, BC, V6T 2A1, Canada; School of Kinesiology, Faculty of Education, University of British Columbia, Vancouver, BC, V6T 1Z1, Canada; Schulich School of Medicine & Dentistry, Western University, London, ON, N6G 2M1, Canada

**Keywords:** racialized, newcomers, chronic disease prevention, heath promotion programs, immigrant, refugee

## Abstract

**Background:** Health promotion programs targeting chronic disease prevention and management (CDPM) are crucial for mitigating the growing the burden of chronic disease and addressing health disparities and preventable risk factors in diverse populations. Racialized newcomers face a disproportionate risk of chronic disease and encounter barriers in accessing healthcare and engaging fully in such programs, hence further compounding health disparities. Engagement is defined as a multifaceted concept that frames involvement through the lens of recruitment, retention, adherence and full participation in an intervention. While culturally tailored programs have demonstrated promise once individuals are engaged, there is a gap in literature synthesizing engagement strategies for these populations in CDPM interventions.

**Objective:** This scoping review aims to synthesize evidence on the engagement strategies used in CDPM interventions for racialized newcomers in Western countries.

**Methods:** We followed Arksey & O’Malley’s (2005) framework along with refinements by Levac et al. (2010), screening 3,898 articles across multiple databases. Data were extracted using the Elicit AI tool along with verification of extraction from research team members. Descriptive analysis summarized findings.

**Results:** Forty-eight studies were included. Most studies focused on Hispanic/Latino and East/Southeast Asian populations in the USA. Common engagement strategies included culturally tailored content, use of community health workers (CHWs), accessible materials, financial incentives, flexibility, and co-creation with community input.

**Conclusion:** Culturally relevant and community-driven strategies, including CHWs, and creative and flexible program formats, are key to engaging racialized newcomers in health promotion programs. Addressing barriers and involving communities in program design can improve participation and outcomes in chronic disease prevention and management.

## Background

Health promotion programs, particularly those focused on chronic disease prevention and management, play a vital role in mitigating population health disparities and improving health outcomes across diverse communities [1]. In recent years, many Western countries, including Canada, France, Switzerland, and the United Kingdom, have seen substantial increases in immigration, leading to a greater diversification of their populations [2]. As a result, newcomers, especially those from racialized (or visible minority) groups, often face significant health barriers upon integration into these new countries [3–5]. Barriers include language and environmental difficulties, cultural differences, lack of social support networks, and experiences of systemic discrimination [6, 7]. These barriers contribute to disparities in health outcomes and hinder the effective engagement of these populations in health promotion programs designed to prevent or manage chronic diseases [8–11]. Such engagement challenges can ultimately limit participation in health promotion programs, exacerbating health inequities and contributing to poorer chronic disease outcomes among racialized newcomer populations [9].

Despite the increasing diversity in many high-income countries, there remains a notable gap in the literature synthesizing how racialized newcomers are engaged in health promotion initiatives focused on chronic disease prevention and management [12, 13]. This gap is particularly concerning given the growing recognition of the importance of culturally relevant and accessible interventions in promoting health equity. Understanding the strategies used to foster engagement, the nature of the health promotion interventions that follow, and the populations they aim to serve is essential for informing future research, shaping effective policies, and improving program development.

### Conceptualizing the Term Newcomer

The term *newcomer* is widely used in policy, practice, and academic contexts, yet its meaning can vary considerably depending on the framework and intent of its use. Particularly in the context of health promotion and chronic disease prevention, this term often lacks consistency in how it is defined and operationalized across studies. Barrick et al. [14] propose *newcomer* to be a more comprehensive and inclusive term compared to various conceptualizes of terms such as immigrant, refugee, asylum seeker, etc. The use of the term newcomer in this review is thus both inclusive and critically reflective. It considers how immigration pathways, settlement experiences, and systemic barriers intersect to shape health outcomes and access to care.

Additionally, while length of time since arrival is often used to categorize individuals as newcomers (e.g., five years or less) [15], this review recognizes that the health and social challenges associated with resettlement and marginalization can persist well beyond this threshold. Consequently, some interventions may engage individuals who have been in the host country for a longer duration of time but still face similar barriers as individuals who arrived in the host country more recently. In this review, *newcomer* is conceptualized as an umbrella term that typically refers to individuals who have recently arrived in a country other than their place of birth. This can include immigrants, permanent residents, refugees, and asylum seekers. We have decided to exclude temporary migrants and workers in this definition in order to acknowledge individuals and groups who intend to or are living in a new country long-term. Given the diversity within newcomer populations, and the structural inequities they often face, a nuanced understanding of this term is critical to addressing the unique challenges faced by this population.

This review focuses specifically on newcomers who are members of racialized and ethnocultural minority groups. These individuals are often subject to systemic inequities due to the intersection of their newcomer status with visible minority identity, which may include African, Caribbean, Latin American, South Asian, East/Southeast Asian, Middle Eastern, and other non-white ethnic backgrounds [15]. It is important to acknowledge that the term racialized emphasizes the social and political processes by which certain groups are ascribed racial meanings, often leading to differential treatment and access to resources, including healthcare [16].

The objective of this scoping review is to provide an overview of the existing literature on health promotion programs, operationalized as chronic disease prevention and management (CDPM) interventions, that target racialized newcomer populations in Western countries.

Specifically, this review sought to answer the question: *What is known from the existing literature about engaging racialized newcomers in health promotion interventions?* To address this, we framed engagement as a multifaceted concept that centers on participant involvement; engagement strategies include those that seek to recruit newcomers into tailored interventions, retain participants, maintain adherence to program protocols and remove barriers to full participation in the intervention. This review synthesizes literature on how newcomer populations are engaged in CDPM programs, including the populations served, the strategies employed to promote engagement, and the challenges or barriers encountered in implementing these interventions. The overall goal of conducting this scoping review is to inform and enhance the design and delivery of future health promotion initiatives for racialized newcomers; to ensure engagement strategies employed are appropriate and effective; and to address current barriers that prevent newcomers from participating in health promotion programs which in turn create subsequent inequities in program deliveries and health outcomes.

## Methods

We followed the methodological framework proposed by Arksey & O’Malley [17] and further refined by Levac et al. [18]. This framework involves a five-stage process: (1) identifying the research question, (2) identifying relevant studies, (3) selecting studies, (4) charting the data, and (5) collating, summarizing, and reporting the results. In line with recommendations by Levac et al. [18], particular attention was paid to enhancing methodological rigor through iterative team discussions at each stage, clarification of inclusion criteria, and an emphasis on both descriptive and analytical synthesis of the findings.

### Identifying Relevant Studies, Search Strategy

The original search was conducted in July 2024 and subsequently updated in March 2025. The databases searched included: Embase (OVID), MEDLINE (OVID), CINAHL (EBSCOhost), and Scopus (Elsevier). The initial search strategy was developed in collaboration with an academic librarian, who conducted preliminary test searches to refine the scope and ensure alignment with the research objectives. The development of the search terms and structure was informed by relevant existing literature, as well as iterative feedback from the research team, which consisted of health promotion researchers and healthcare professionals. The search strategy included terms related to key concepts such as health promotion programs, newcomers, and chronic disease prevention and management. The final set of search terms and combinations was reviewed and approved by the research team to ensure comprehensiveness and relevance. The search terms are provided in Table 1, and the full Medline search strategy is provided in Appendix 1.

**Table I.** Examples of search terms.

| Newcomer | Health Promotion Program |
| --- | --- |
| “Emigrant”, “Immigrant”,<br>“Immigration”, “Emigration”,<br>“Refugee”, “Refugees”, “Asylum<br>Seeker”, “Asylum Seekers”, “Asylee”,<br>“Displaced Person”, “Displaced<br>Persons”, “Incomer”, “In Comer”,<br>“Newcomer”, “New Comer”, “Migrant”,<br>“Migrants”, “Resettler”, “Resettlers”. | “Lifestyle Intervention”, “Chronic Disease Management Program”,<br>“Chronic Disease Prevention Program”, “Diet”, “Behavior Change”,<br>“Behaviour Change”, “Weight Loss Program”, “Healthy Living Program”,<br>“Healthy Lifestyle Program”, “Health Education”, “Medical Education”,<br>“Physical Education and Training”, “Education”, “Health Promotion”,<br>“Exercise”, “Community Networks”, “Social Support”, “Health Policy”,<br>“Health Promotion Program”, “Chronic Disease Prevention Management<br>Program”, “Weight Loss Intervention”, “Healthy Eating Promotion<br>Campaign”, “Community-Based Program”, “Community Led Health<br>Promotion”, “Community Cardiovascular Health Promotion Program”,<br>“Physical Activity”, “Nutrition Intervention”. |

### Study Selection

The initial search yielded 3,898 articles. After removing duplicates, 2,819 titles and abstracts were screened by pairs of independent reviewers. Screening was conducted by four members of the research team (MK, PA, AT, JO) using established inclusion and exclusion criteria.

Following this stage, 191 articles met the criteria for full-text review. Each full-text article was assessed independently by pairs of reviewers. Any conflicts during the screening process were resolved by including a third reviewer.

### Inclusion and Exclusion Criteria

See Table II for inclusion and exclusion criteria utilized.

**Table II.** Inclusion and exclusion criteria.

| Selection criteria | Inclusion criteria | Exclusion criteria |
| --- | --- | --- |
| Study design | Empirical research articles, published at any time | -Non-empirical studies (e.g., commentaries, opinion pieces, editorials, or theoretical papers)<br>-Reviews (systematic, scoping, narrative)<br>-Protocols without reported outcomes<br>- Feasibility, pilot, or acceptability studies without reported intervention/ effectiveness outcomes<br>-Case studies, conference abstracts, and dissertations (if applicable) |
| Population | -Adults or family-based interventions<br>-Racialized newcomers or immigrants residing in Westernized countries (e.g., Canada, UK, USA, New Zealand, Australia)<br>- Newcomer categories include: refugees, asylum seekers, and immigrants | -Temporary settlers such as: temporary foreign workers, migrants, and international students<br>-Youth<br>-Non-racialized newcomers/immigrants |
| Language of publication restrictions | English language | Languages other than English |
| Intervention design | -Interventions focused on health promotion for chronic disease prevention or management (Chronic Disease Prevention and Management – CDPM programs)<br>-Target modifiable risk factors such as physical activity, sedentary behavior, and healthy eating<br>-Intervention/effectiveness studies that report on outcomes |  |

#### Inclusion criteria

For the purposes of this review, chronic disease prevention and management (CDPM) programs were defined according to the definition outlined by Gavarkovs, Burke & Petrella [19]. Chronic disease prevention programs were operationalized as interventions aimed at modifying lifestyle-related risk factors, such as diet, physical activity, or weight management, to prevent the onset of chronic illness [19]. In contrast, chronic disease management programs refer to interventions designed to support individuals already diagnosed with a chronic disease in reducing the burden of illness and improving overall health outcomes [19]. This dual focus on prevention and management reflects the broad scope of health promotion initiatives relevant to newcomer populations and provides a consistent foundation for study inclusion.

#### Exclusion criteria

Studies were excluded if they did not meet the criteria outlined in Table II. Specifically, we excluded non-empirical articles (e.g., commentaries), studies lacking intervention effectiveness data, and studies involving non-racialized populations, youth, or temporary residents such as international students, temporary migrants, or temporary workers. Interventions that did not focus on modifiable risk factors (e.g., physical activity or diet) related to chronic disease prevention or management were also excluded. When studies used multiple terms to describe their population (e.g. both ‘migrants’ and ‘immigrants’), the research team made inclusion decisions by consensus, based on the authors’ use of terminology and investigation of the population characteristics described [20].

### Data Extraction

We used the Elicit AI tool [21] to support the data charting and extraction stages of the review. The data extraction form was first created at the initial protocol stage of this review and included information such as engagement strategies, recruitment strategies, and key findings. Once study screening was complete, the first author (MK) piloted this form with 3 studies to ensure all necessary data was captured appropriately [22]. We then used Elicit to facilitate the extraction and organization of key data elements from 3 included studies. The research team continued to refine this extraction to ensure all additional and necessary data was captured such as chronic disease/condition, racialized group, recruitment strategies, engagement strategies, suggested implications, and key learnings related to newcomer engagement. The extraction was then completed with all remaining included studies. All outputs were then reviewed and verified by independent pairs of the research team using an Excel spreadsheet to ensure accuracy and relevance of extraction.

### Data Analysis

We used descriptive analysis to synthesize and summarize data extracted from the included studies. This approach allowed us to systematically collate the characteristics and findings of the studies. As described by Peters et al. [22], including a descriptive analysis in this scoping review provides an overview of the scope and trends within the literature, while focusing on study characteristics, engagement strategies, and intervention.

## Results

A total of 48 articles met the inclusion criteria for this review. The study selection process, including detailed reasons for the exclusion of full-text articles, is illustrated in Figure 1.

**Figure 1.**
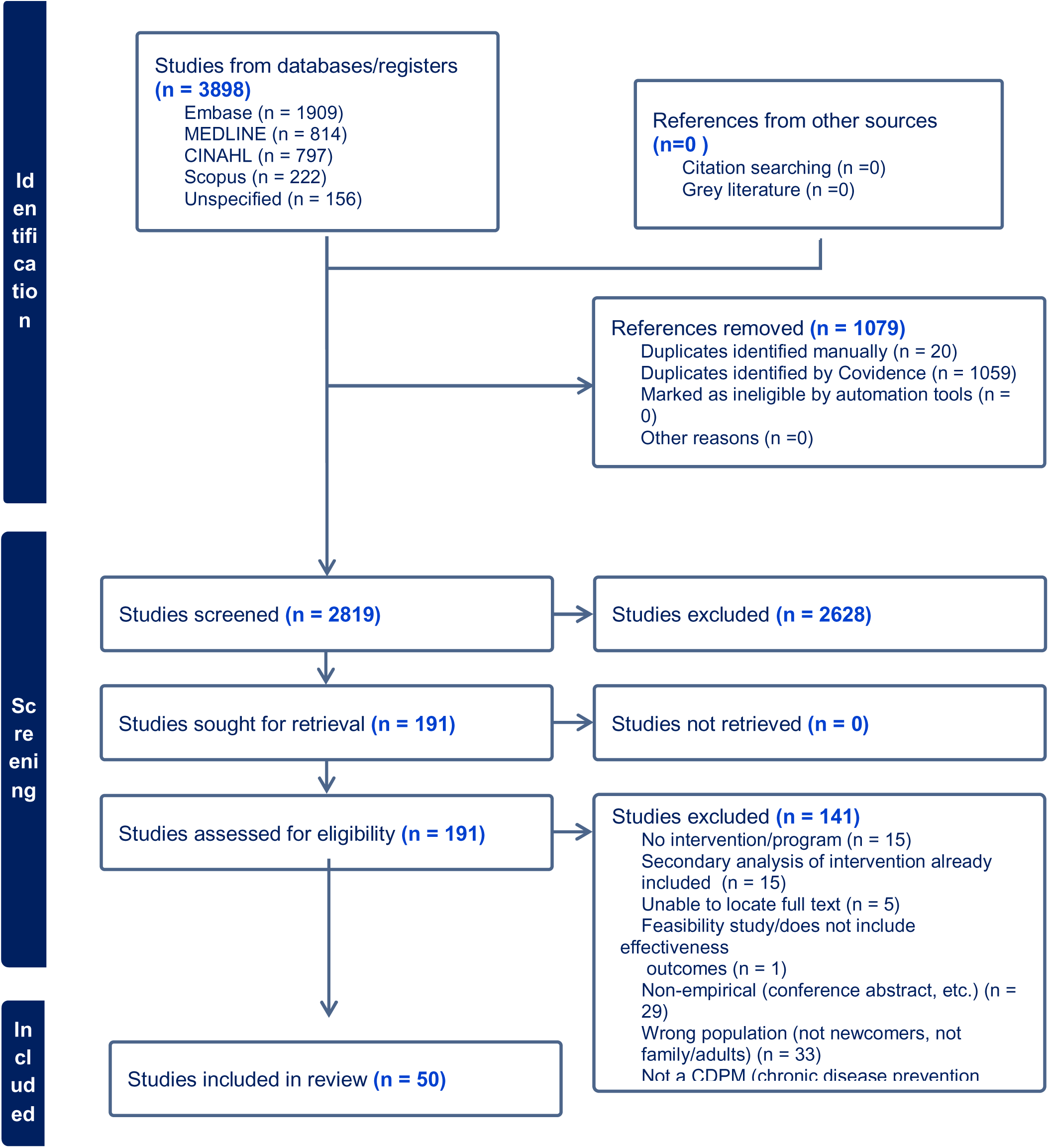
PRISMA flow diagram for included studies.

### Description of Included Studies

This review captured 48 diverse health promotion interventions that engaged racialized newcomers. Among the 48 included studies, the majority were conducted in the United States (n=37), followed by Italy (n=2), Canada (n=2), other European countries (n=6), and Australia (n=1). Most studies included individuals from Hispanic/Latino (n=13) and East/Southeast Asian groups (n=13). Seven studies (14%) included West/Central Asian populations, five (10%) included South Asian, and four (8%) focused on African populations. Additionally, six studies (13%) included adults from more than one of these racialized groups. See Table III.

**Table III.**
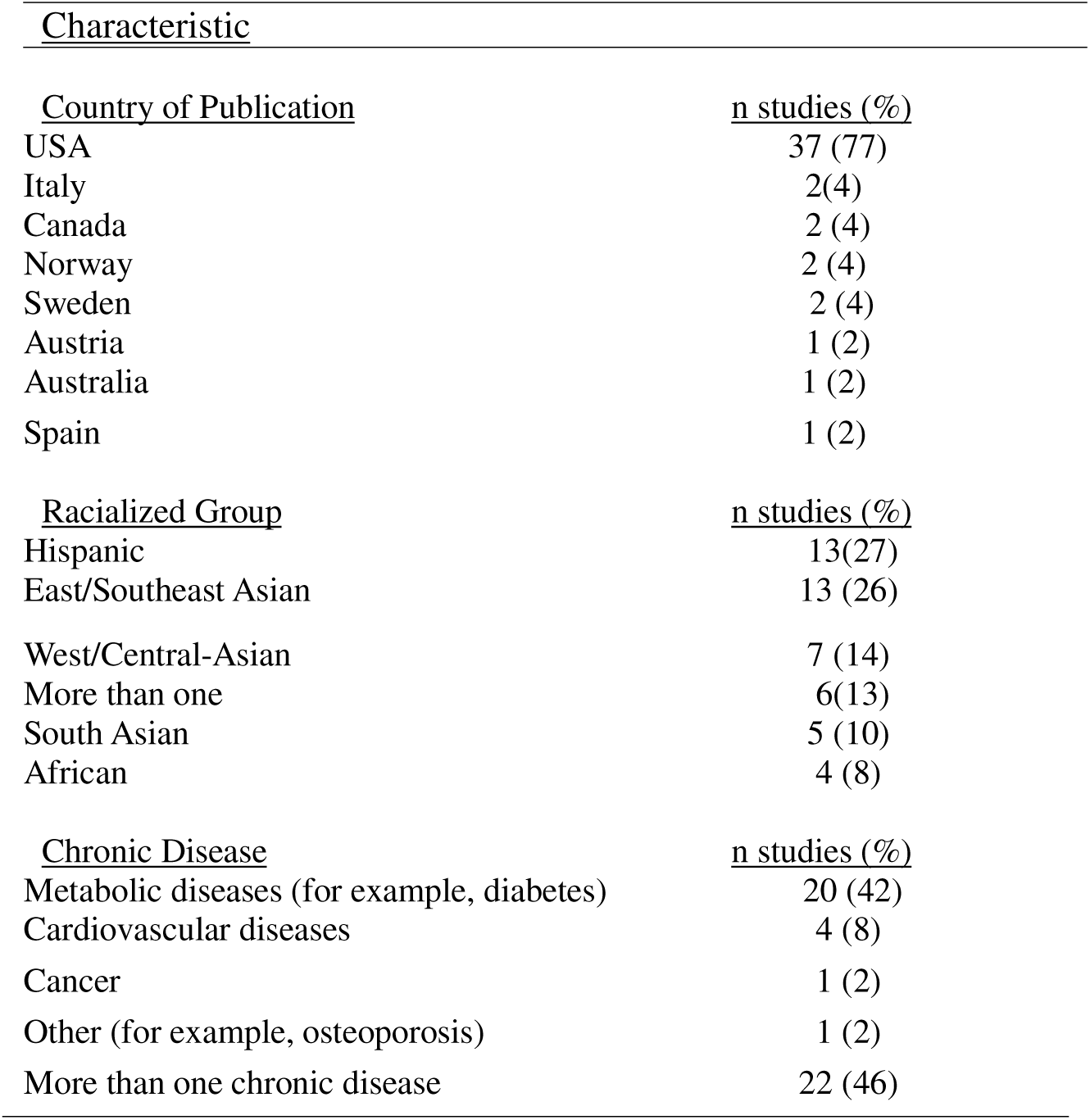
Description of included studies (n=48).

| <u>Characteristic</u> |  |
| --- | --- |
| <u>Country of Publication</u> | <u>n studies (%)</u> |
| USA | 37 (77) |
| Italy | 2(4) |
| Canada | 2 (4) |
| Norway | 2 (4) |
| Sweden | 2 (4) |
| Austria | 1 (2) |
| Australia | 1 (2) |
| Spain | 1 (2) |
| <u>Racialized Group</u> | <u>n studies (%)</u> |
| Hispanic | 13(27) |
| East/Southeast Asian | 13 (26) |
| West/Central-Asian | 7 (14) |
| More than one | 6(13) |
| South Asian | 5 (10) |
| African | 4 (8) |
| <u>Chronic Disease</u> | <u>n studies (%)</u> |
| Metabolic diseases (for example, diabetes) | 20 (42) |
| Cardiovascular diseases | 4 (8) |
| Cancer | 1 (2) |
| Other (for example, osteoporosis) | 1 (2) |
| More than one chronic disease | 22 (46) |

Most interventions were delivered in community settings (such as health centres, faith-based spaces, or local immigrant-serving organizations). The majority of studies focused on chronic disease prevention (rather than chronic disease management), particularly related to diabetes and cardiovascular disease. Almost all studies focused on adults (n= 44), while four studies focused on families [23–26].

### Engagement Strategies

Across the included studies, culturally-tailored strategies, use of community health workers, accessible language materials, community partnerships, providing incentives, and flexible scheduling emerged as common engagement mechanisms. These strategies were aimed to support ongoing retention, adherence, and active participation throughout the intervention period. Programs that embedded community-based participatory research (CBPR) principles reported higher adherence and retention. An overview of key findings of included studies are summarized in Table IV.

**Table IV.**
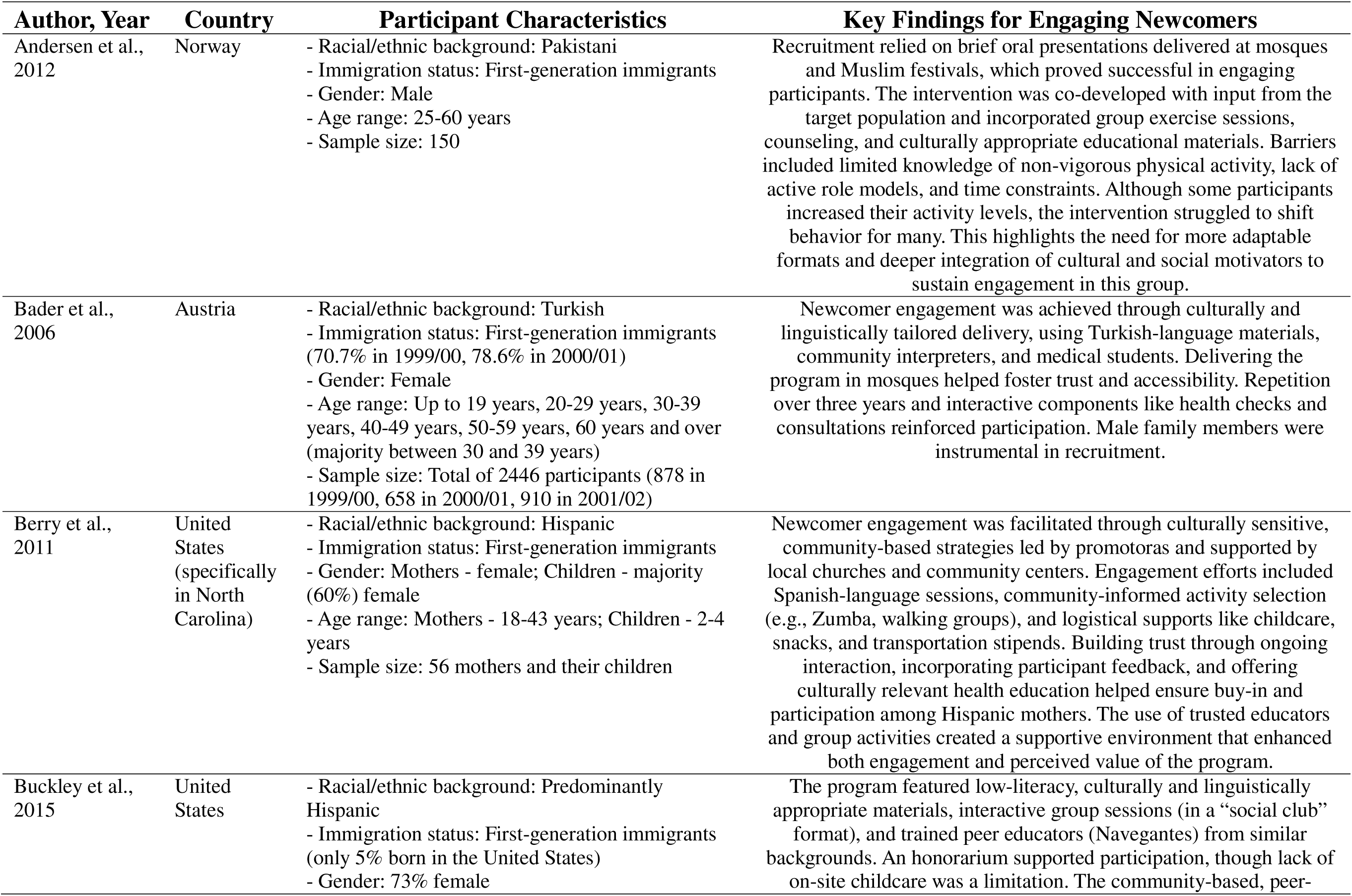
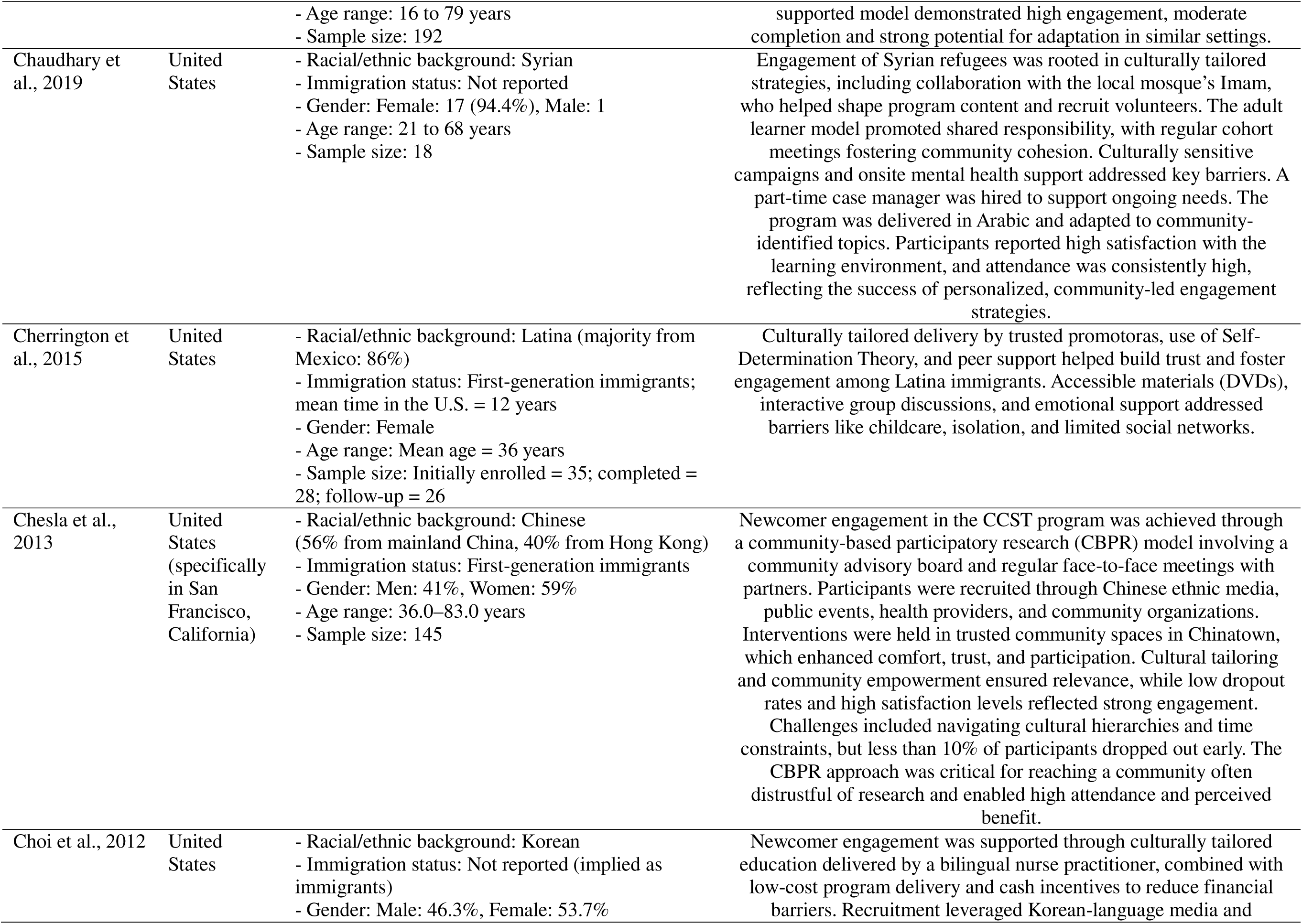
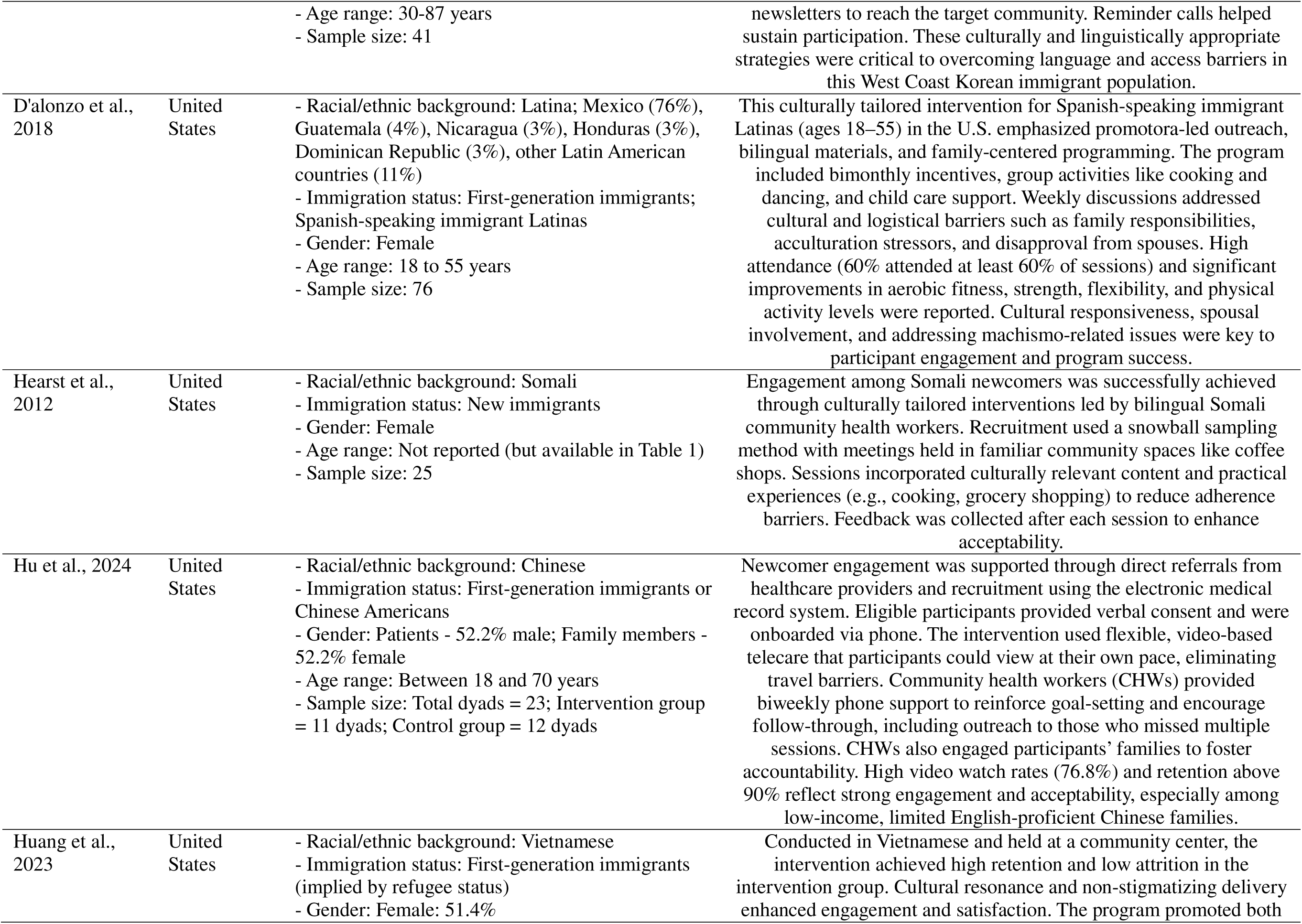
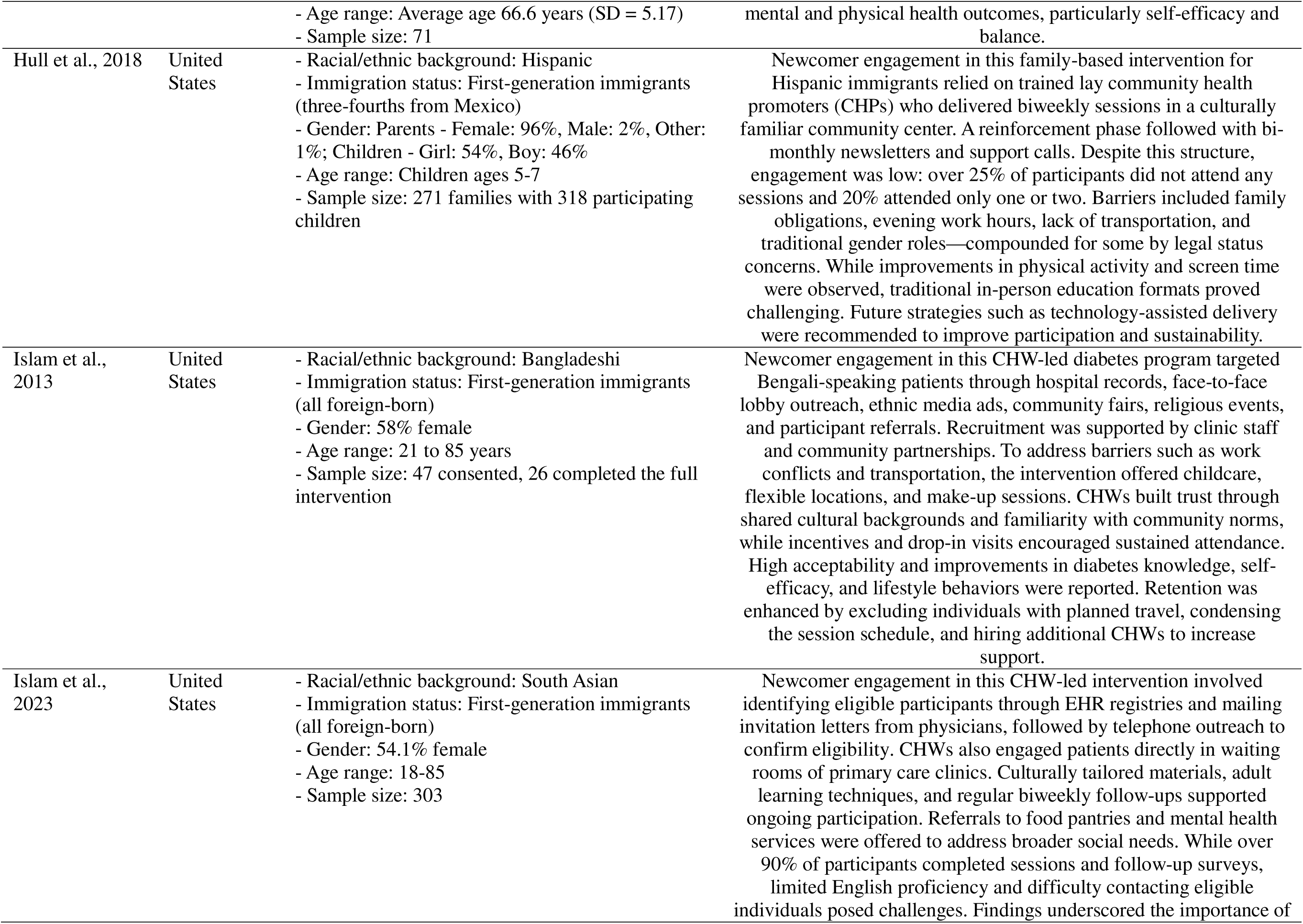
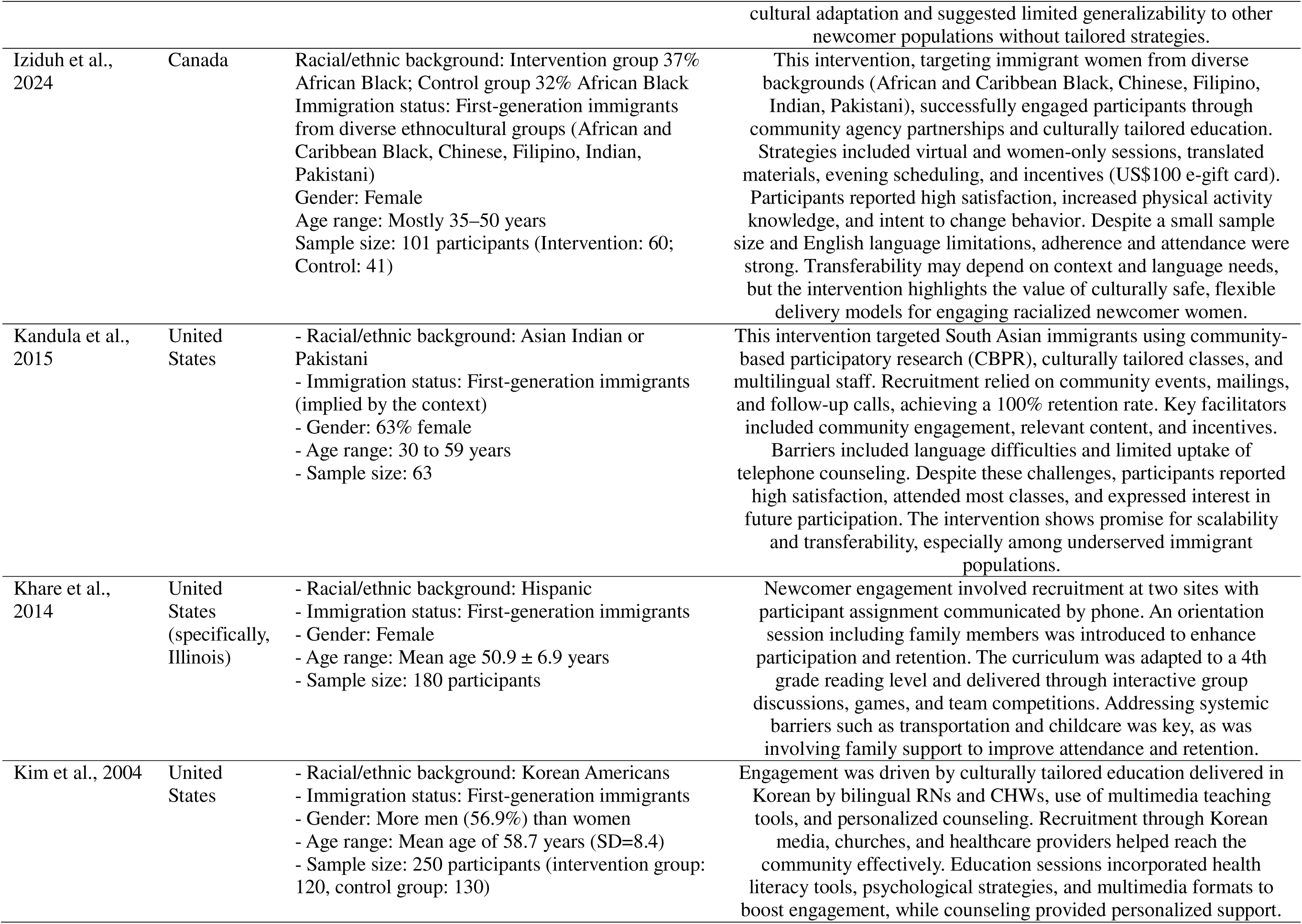
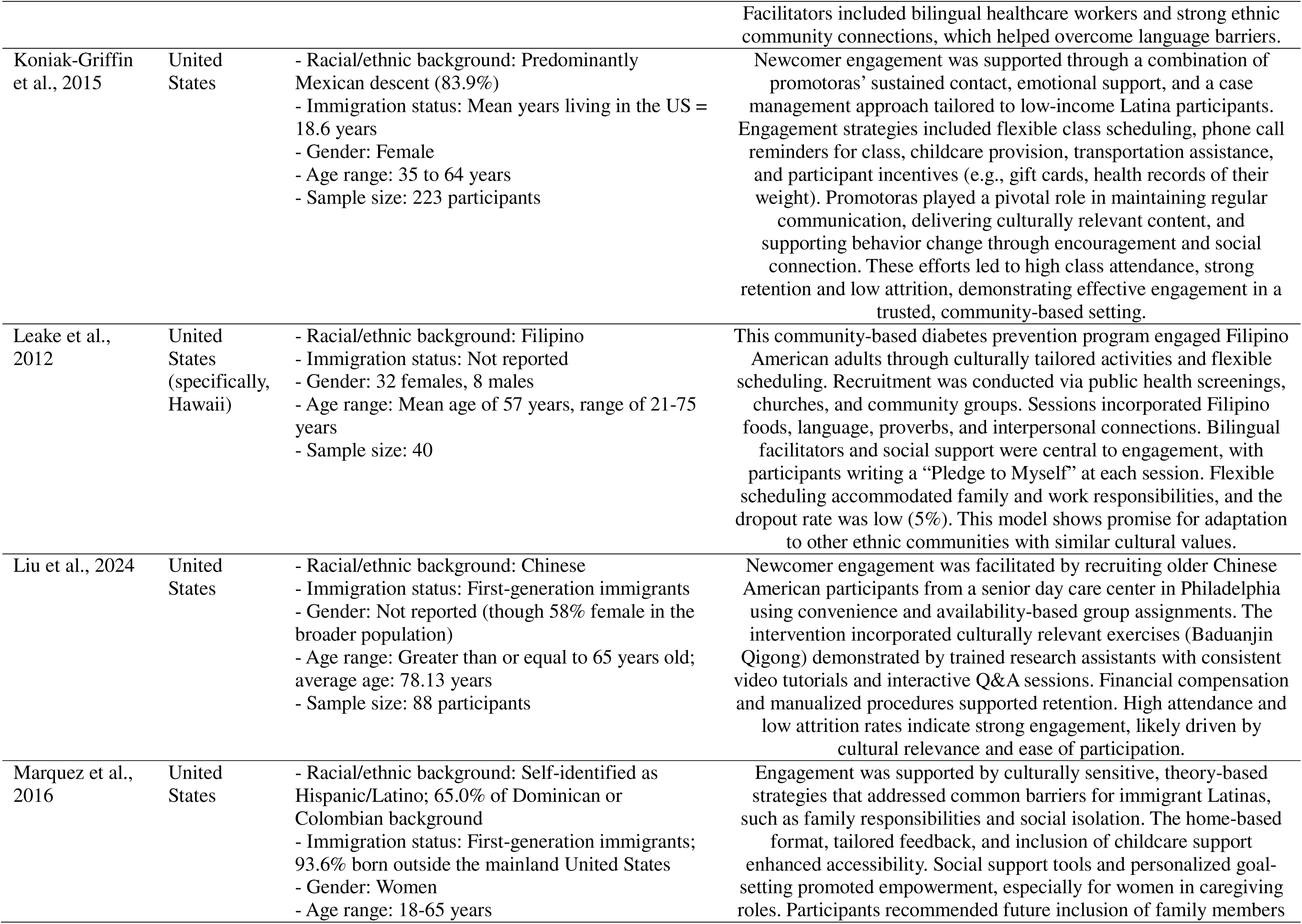
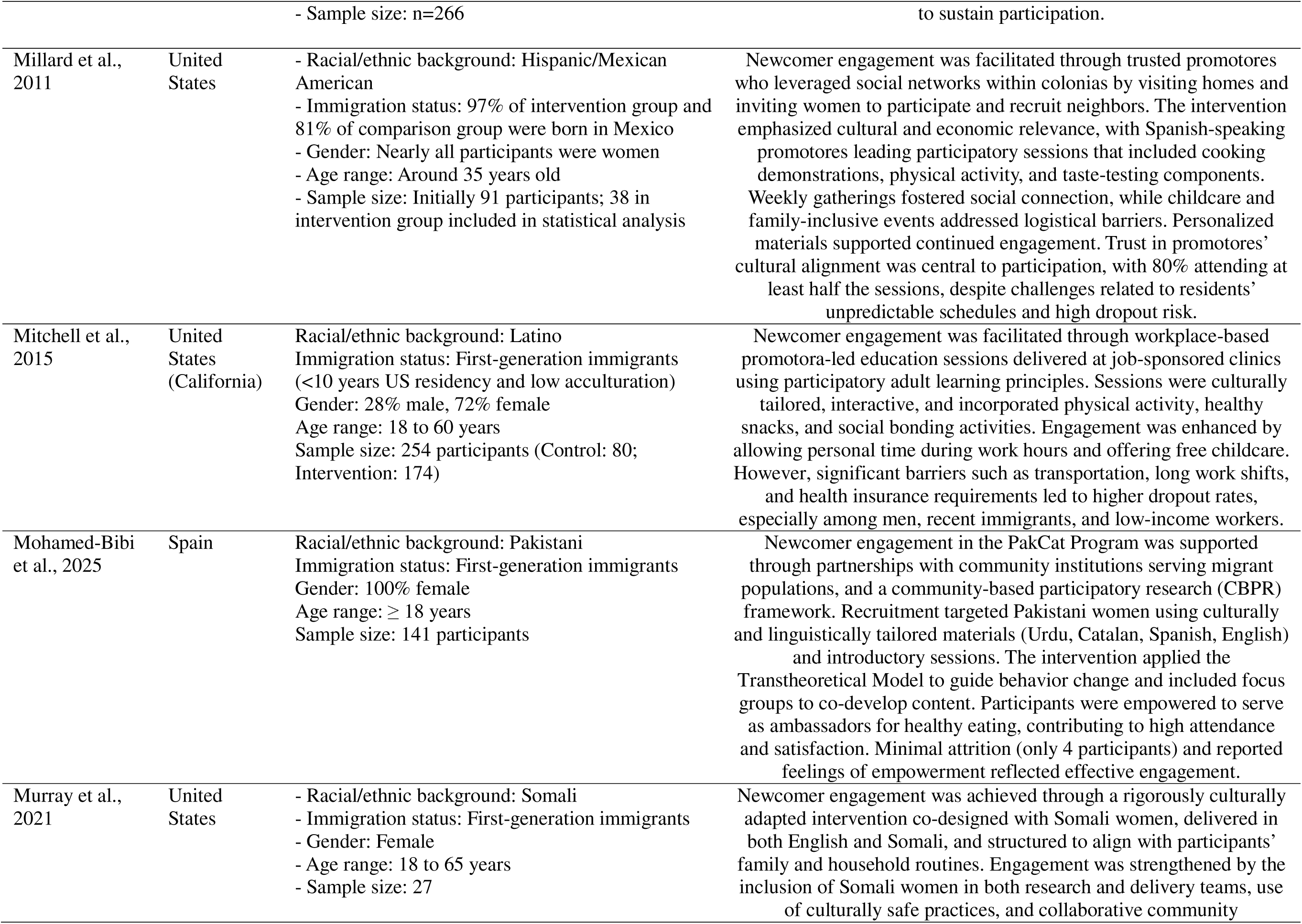
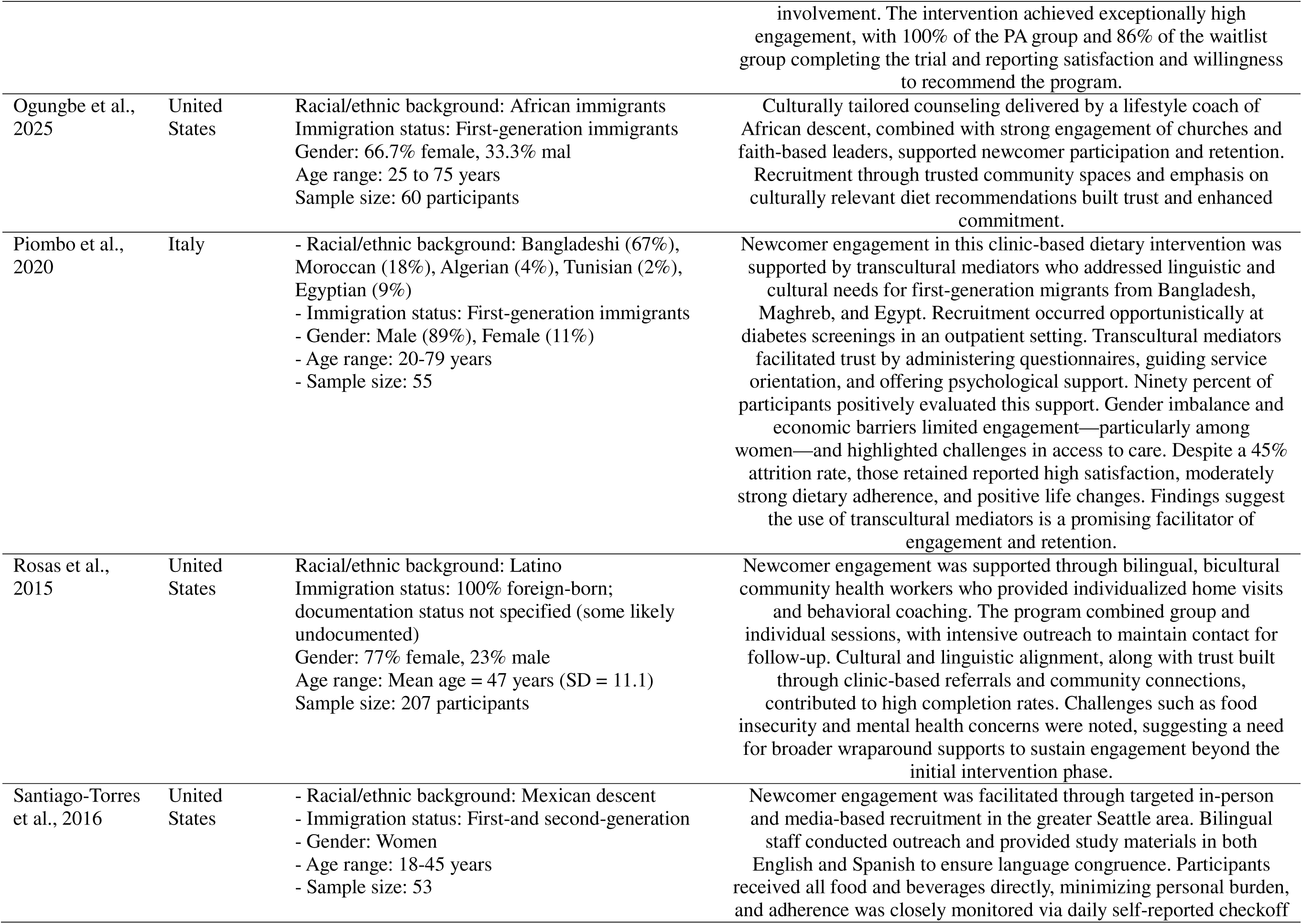
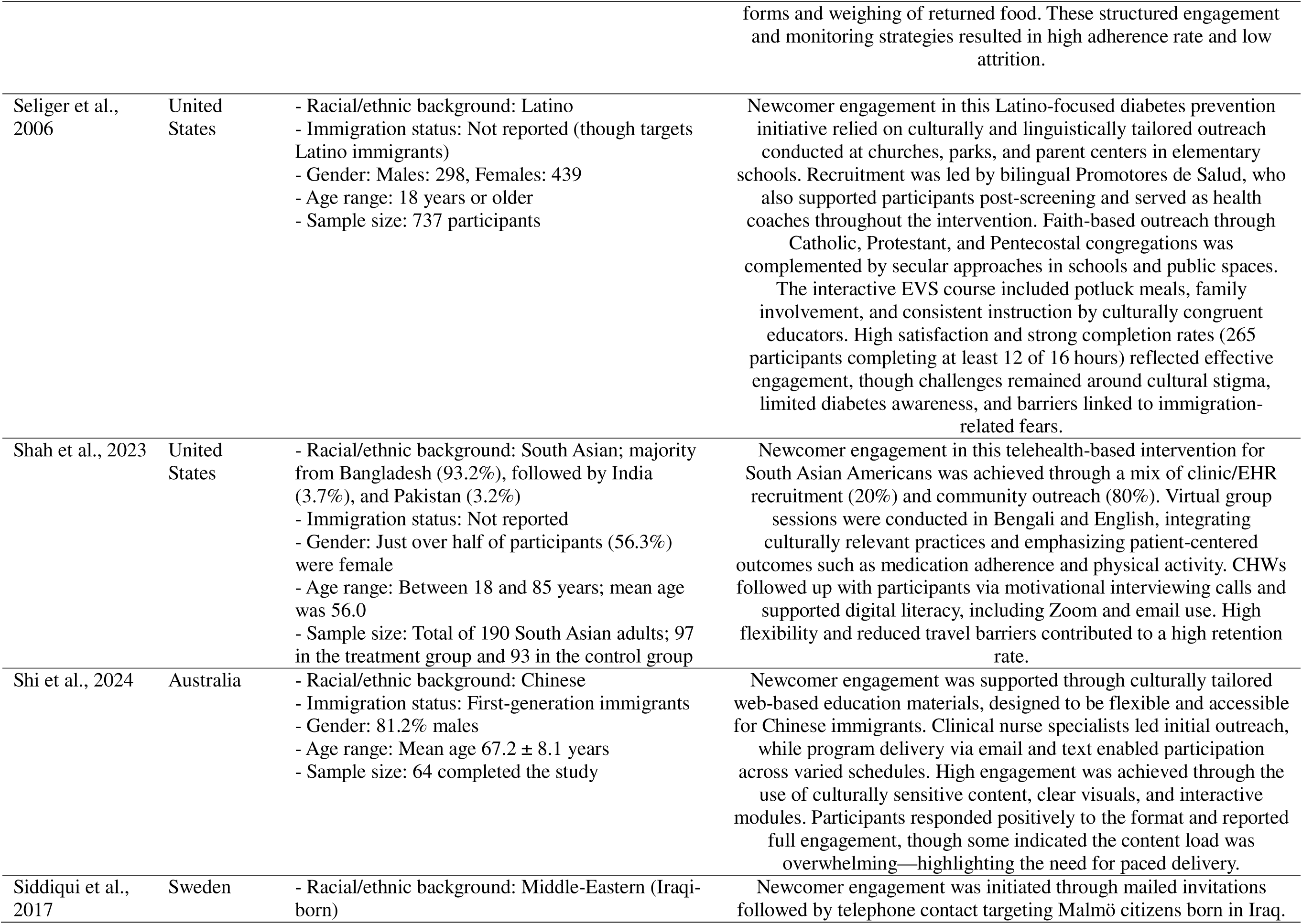
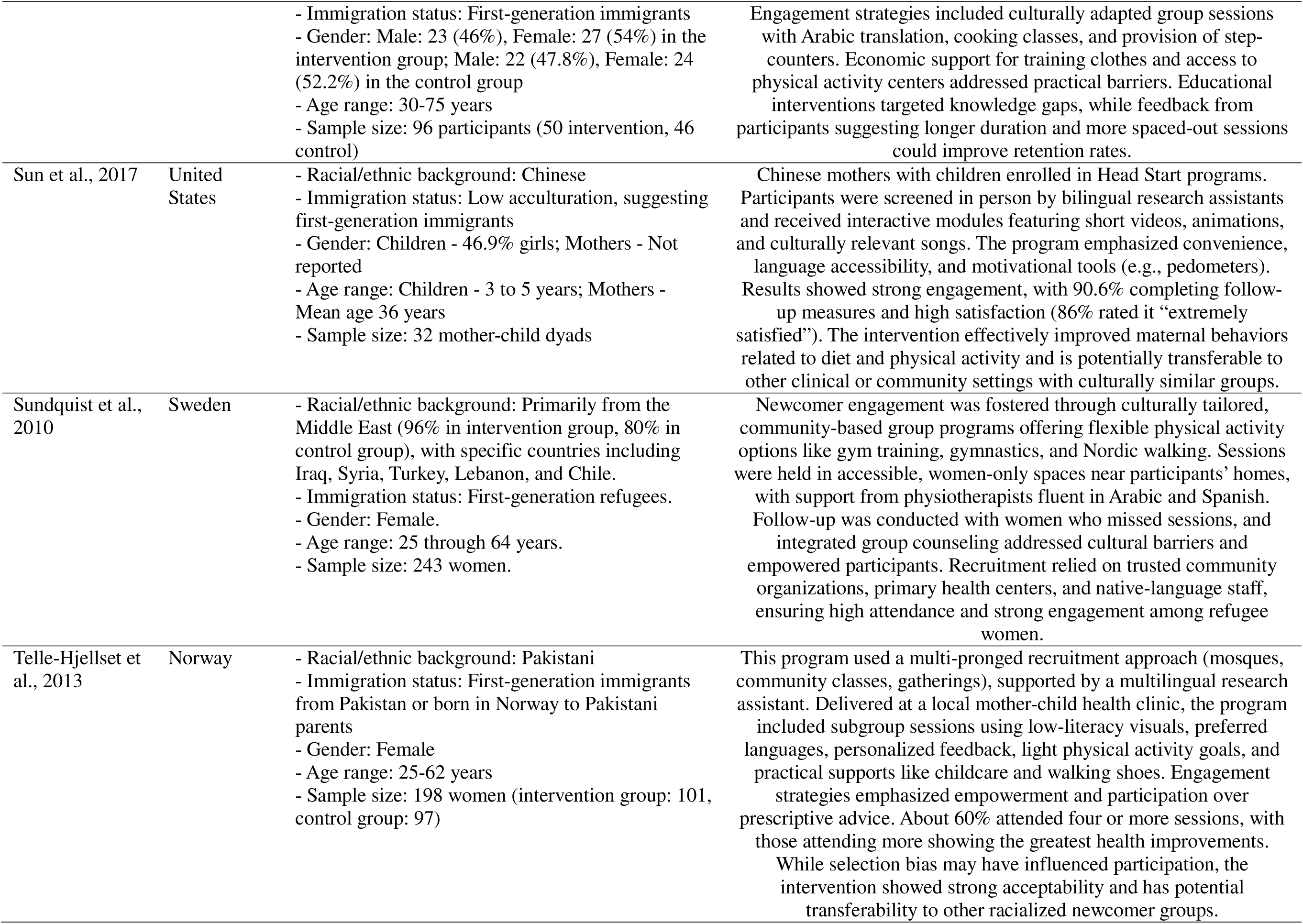
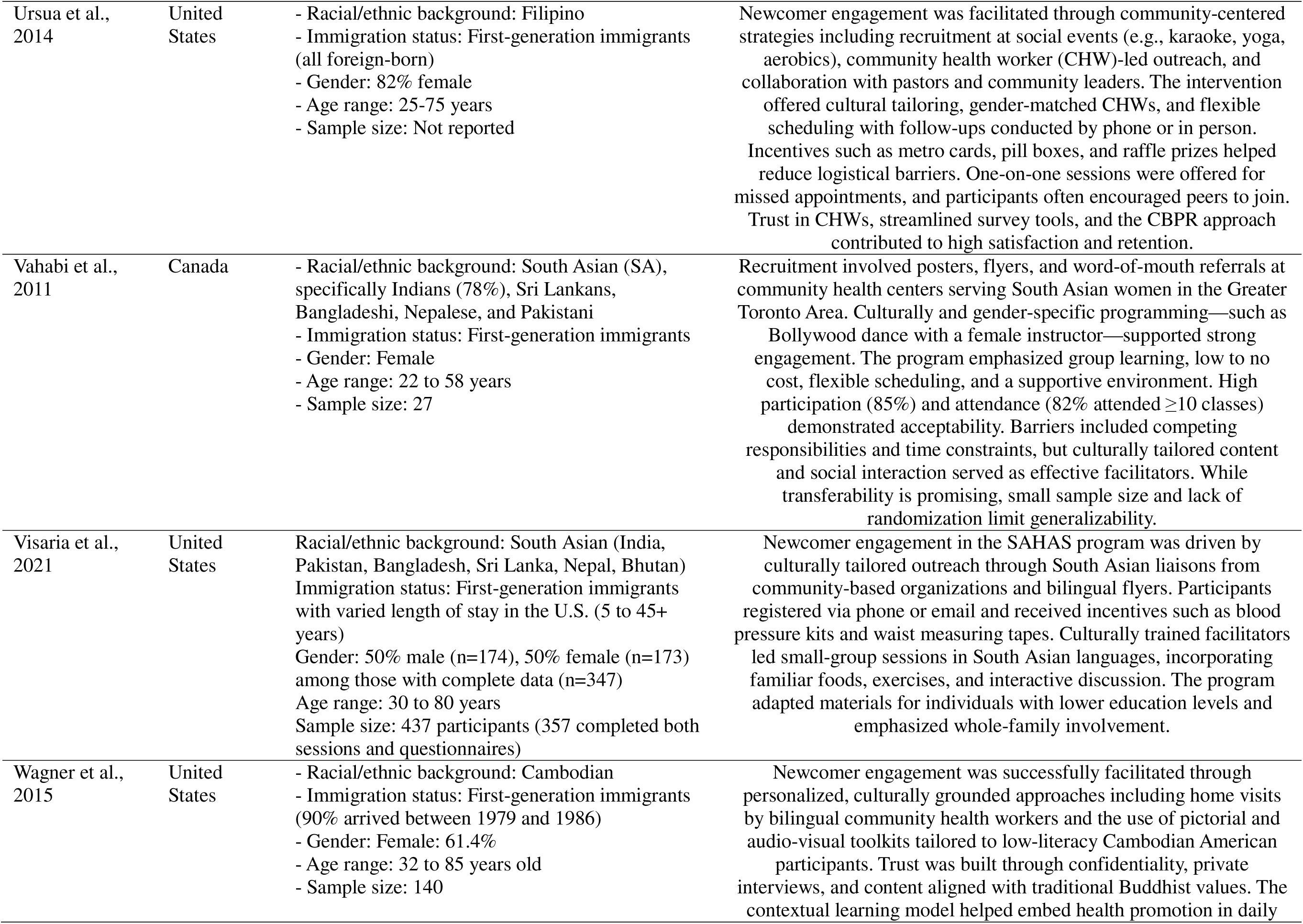
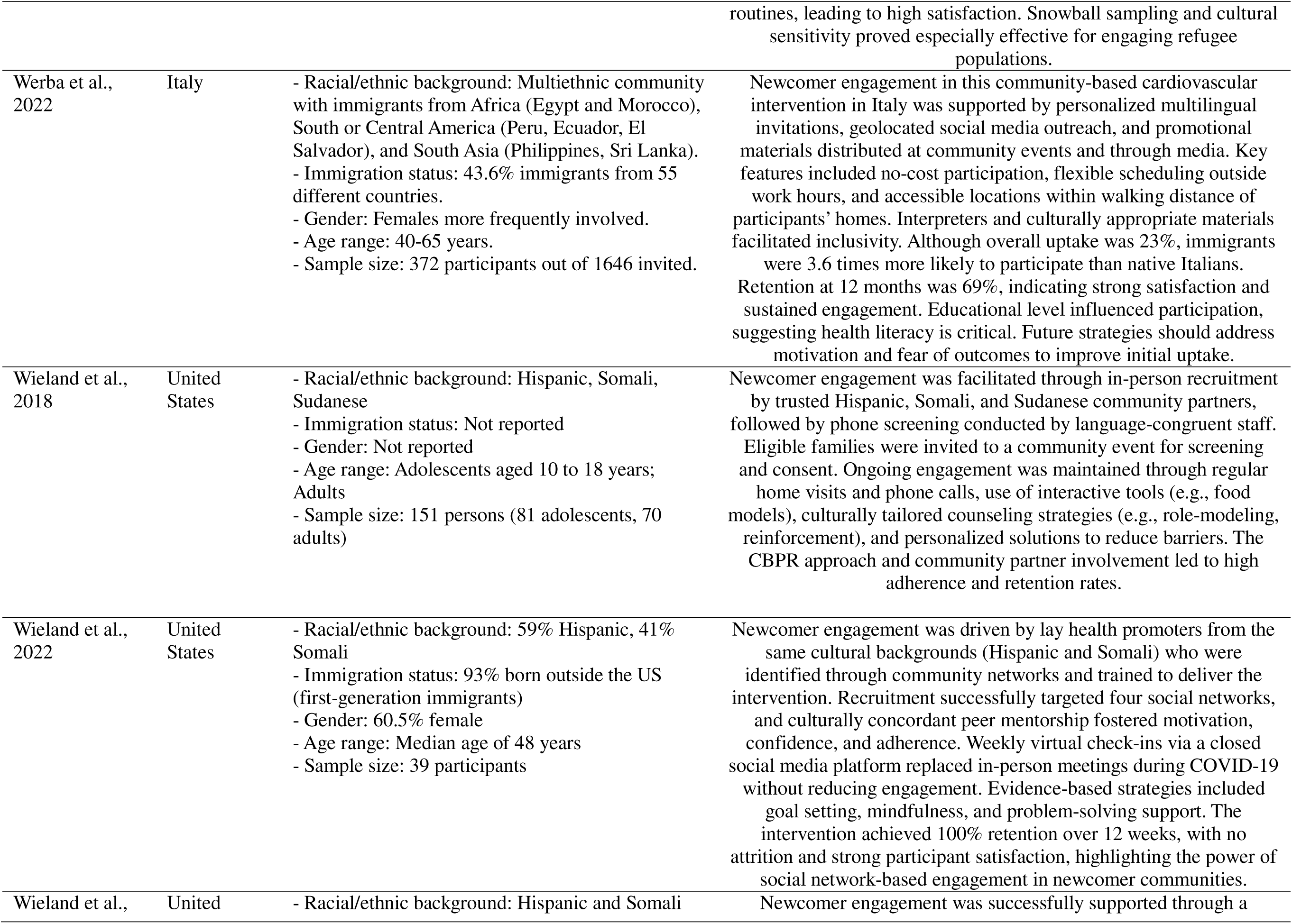
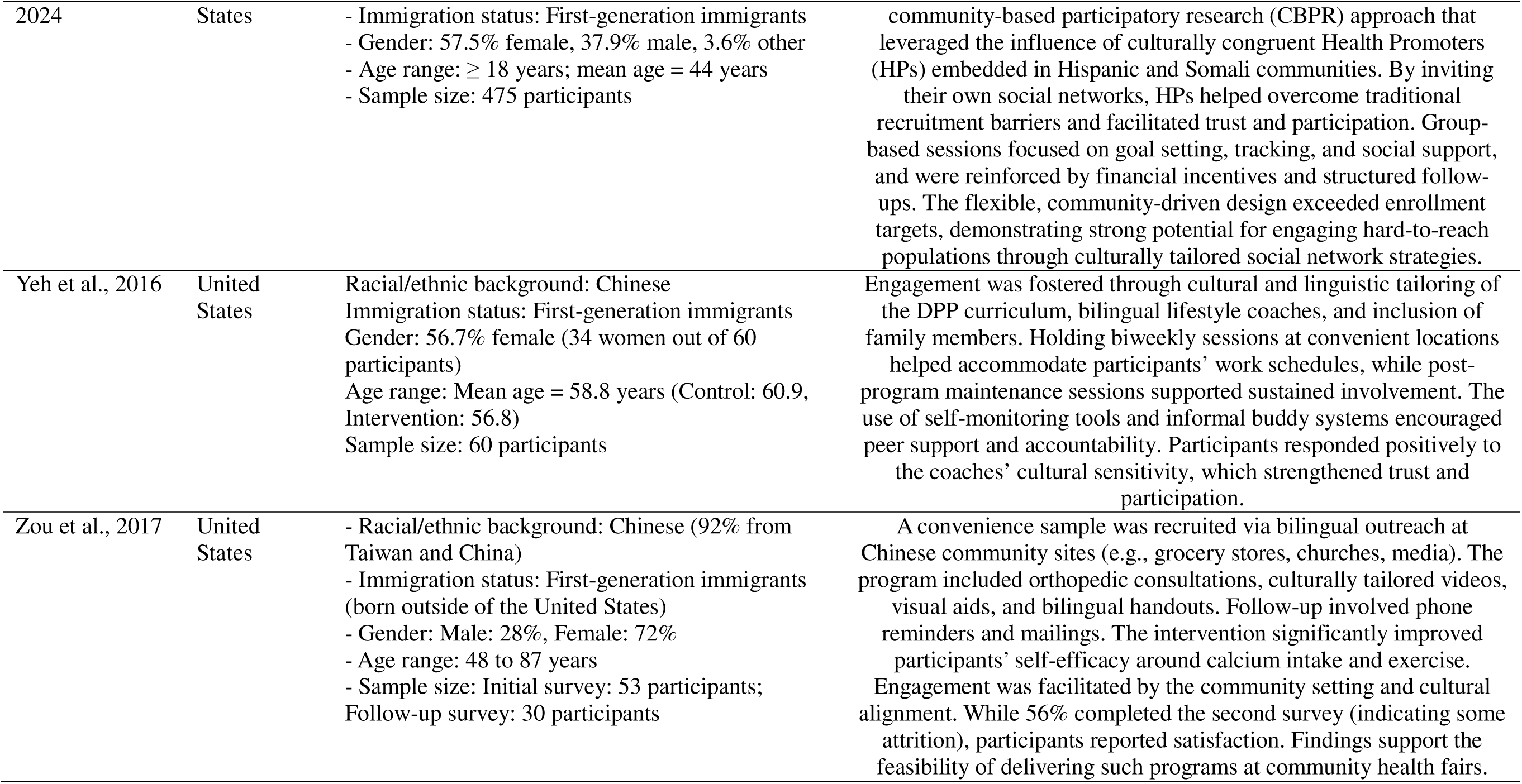
Description of included studies.

#### Recruitment Strategies

Several studies highlighted the use of community health workers (CHWs) and other trusted community figures as key recruitment strategies to reach racialized newcomer populations. Commonly, CHWs were critical for establishing rapport and trust within communities, leveraging their deep cultural connections and shared sociodemographic characteristics. For example, participants were recruited through informal settings such as coffee shop meetings, community gatherings, or local centers frequented by newcomers [23, 27]. These environments facilitated open conversations about health programs and encouraged involvement, especially in communities where healthcare access was limited. Other studies integrated more formal recruitment methods within healthcare settings, such as utilizing medical centers and electronic medical records to identify and contact eligible participants, or by having CHWs recruit patients in primary care waiting rooms [28–35]. In these cases, healthcare providers or CHWs helped to bridge the gap between the medical system and underserved populations by identifying individuals at risk for chronic diseases or who could benefit from health promotion programs.

Some studies also suggested tailoring recruitment efforts made by same-gendered individuals to promote comfortability and cultural appropriateness [48, 65]. Lastly, other studies utilized printed flyers mailed to homes and posted at grocery stores, churches, and community centers to recruit participants [38, 39]. These recruitment strategies, particularly those combining both formal and informal methods, ensured broad reach and helped recruit populations that might otherwise remain disconnected from health promotion opportunities.

#### Building Trust in the Community

Studies often leveraged existing and strong community ties to mobilize community engagement and build social support. [24, 27, 34, 36, 38–55]. This included having the research team build trust with key community members prior to and during the program, which included attending mosques and other community gatherings [27, 45]. This also included attending social activities such as aerobics or line dancing events [48]. Building strong ties with social service agencies was also key, as these agencies provide employment and housing support, and/or language classes for newcomers [42]. One study empowered ‘health promoters’ or opinion leaders in the community to recruit participants through their own social networks [47], whereas other studies created deep ties to faith-based organizations and leaders to build trust and sustainability [48, 49, 53]. Offering introductory orientation sessions to explain the program was also important in fostering trust and promoting engagement [52].

#### Financial Incentives

Almost half of the included studies (n=21; 42%) engaged participants through various financial incentives [23, 25, 31, 34, 36, 38, 42, 50, 51, 53, 54, 56–65]. These incentives were often tailored to reduce structural barriers such as transportation and cost, or to acknowledge participants’ time and commitment. Examples of these incentives included providing the intervention for free, reimbursing childcare [60], providing food/meals to participants [25, 57, 61, 62, 65], gift cards of $20-100 value for attendance and participation [34, 54, 56, 59, 60, 64], and bus/metro passes to reduce transportation barriers [31]. Studies also provided fitness clothes and access to fitness facilities [40, 63].

#### Flexibility and Accessibility

Studies (n=14; 29%) explicitly implemented flexible scheduling and accessible delivery methods to support engagement in health promotion intervention [30, 36, 42, 50]. In other cases, interventions were held in familiar and easily accessible and walkable locations, such as neighborhood parks, which made participation more convenient [30, 41, 64].

Flexibility was further supported through strategies like rescheduling sessions, offering makeup classes, allowing participants to attend sessions in any order and to decide on their own class time [41]. These approaches helped accommodate the diverse schedules and commitments of participants, such as work or family responsibilities [40, 41]. Additionally, several interventions provided childcare services to address the needs of parents and caregivers [27, 37, 43, 50, 59, 61–63]. Bi-weekly sessions (once every two weeks) were another successful strategy, as they offered a balance between maintaining participant engagement and accommodating participants’ busy lives [35]. Monthly maintenance check-ins were also used to keep participants on track while allowing them to balance their personal schedules. These strategies not only enhanced accessibility but also helped reduce barriers to continued participation, ultimately improving the effectiveness of chronic disease prevention and management interventions [35].

#### Community Health Workers

Twenty (40%) of the included studies utilized CHWs or *promotoras* to improve program adherence and retention [23, 26, 29, 31, 36, 37, 43, 43, 44, 48, 50, 50, 51, 56, 59, 61, 62, 68–70].

Islam et al. defined CHWs as health professionals who “play an integral role in disseminating effective health interventions through their deep knowledge and connection to communities, cultivated through shared sociodemographic or cultural characteristics and trusted relationships” [33, p. 206]. In these CHW-based interventions, CHWs engaged participants in goal-setting, attending grocery store visits, and delivering encouragement and follow-up phone calls and home visits [29, 66].

#### Language and Cultural Components

All studies (n=48) included language and culturally-appropriate materials as a way to reduce barriers and facilitate participation. Language accessibility was key in enhancing engagement, with several interventions adapting curriculum to appropriate reading levels [grade 4-5; 45, 46], offering bilingual resources, or employing translators to ensure inclusivity and effective communication [54, 63]. Additionally, cultural tailoring in the form of relevant examples, visual aids, and culturally familiar symbols helped bridge cultural gaps and foster a sense of belonging among participants.

One study colorfully decorated their program atmosphere to reflect festive gatherings [41], while other studies held a large shared family meal at project end [59] or at the end of each session [40]. Such environmental adjustments not only made the program feel more welcoming but also encouraged a sense of ownership and pride in the process [41]. Cooking classes were also a helpful strategy to imbed cultural value in programs [23, 63], along with integrating dance as a form of physical activity [33, 42, 58]. These culturally rooted strategies helped participants feel understood and valued, which in turn supported greater engagement through improved adherence and retention, throughout the interventions.

#### Co-creation

Studies (n=11; 23%) included in this review emphasized the need for continued co-creation when engaging newcomers in health promotion interventions. Main approaches to this were through the use of community-based participatory research (CBPR) frameworks, which is an approach that involves equitable collaboration between researchers and community members in all phases of the research process [36, 43, 46–48, 51, 52], and other co-creative processes [25, 67]. Studies completed focus groups with community members prior to program design [43] or continued to involve community members throughout program delivery [67]. Integrating critical religious and spiritual components was also key in the co-creation process [66].

#### Individual Counselling

Studies (n=11, 23%) highlighted the importance of assigning individual counselors, including community health workers and healthcare professionals such as RNs, medical students, or community members, to improve attendance and adherence. These counselors played a key role in fostering personalized engagement by conducting targeted, realistic goal-setting with participants and following up through phone calls [29, 31, 43, 44]. This individualized approach helped ensure that participants remained committed to the program, while also providing the support needed to overcome challenges, ultimately enhancing overall program adherence [25, 32, 43, 45, 68, 69]. Additionally, one study found it helpful to involve transcultural mediators (administrators) when administering questionnaires and surveys to participants [20]. When working with individuals who have experienced trauma, it was also important to offer services such as on-site psychologists for mental health discussions if needed [52].

#### Interactive Approaches

Several studies (n=18; 38%) utilized digital and interactive approaches to enhance engagement. Virtual delivery modes were utilized in some interventions to enhance accessibility, particularly for participants with limited mobility or those facing transportation challenges. Some were entirely digital and web-based [28] and others provided educational video tutorials [24, 32, 70], or were delivered in-home using printed materials [60, 66]. Studies provided participants tools such as Bluetooth-enabled blood pressure monitors [49], measuring kits [45, 50], or pedometers [24, 60]. Another study engaging mother-child dyads assisted participants in creating walking groups and attending park field trips during the program as a way to keep participants engaged [58]. Bilingual “tool kits”, which included pictures and spoken CDs were used to assist participants of lower literacy [66], while another study utilized a ‘buddy system’ to create informal support amongst participants [35]. Additionally, some studies improved engagement by creating a family-based atmosphere [53, 55]. To do this, they welcomed family members to attend sessions and created teams and games to keep participants engaged.

#### Additional Key Learnings

Some studies suggested having gender-separated environments during implementation [30, 33, 42]. As well, specific cultural and religious components were critical to be aware of, as one study ensured proper male and female restricted areas in mosques were respected during the recruitment and implementation processes [45]. Work commitments and transportation continued to be barriers to participant involvement [65], and some studies suggested financial compensation for time missed at work and spaced out sessions would improve retention [63].

Finally, empowering change rather than offering prescriptive advice was shown to improve engagement [27]. This was accomplished in one study by having participants write a “pledge to myself” at the end of each session [41] or by creating weekly promises [62].

## Discussion

### The State of the Literature

The papers included in this review span several contexts and present a variety of approaches to engaging racialized newcomers in CDPM interventions. While a range of settings and intervention styles are explored, the majority of the studies included were conducted in the United States (n=37; 77%) and more than half of the interventions were tailored to Hispanic and East/Southeast Asian newcomer populations; the newcomer experience may be different depending on population and setting, underscoring the need for continued research on CDPM interventions with diverse newcomer communities across Western countries.

### Implications for Future Research and Policy

At a time when many countries are seeing increased levels of immigration, and racialized newcomer populations are experiencing barriers to health program participation, increased risk of chronic diseases, and decreased health outcomes, the papers included in this review provide promising strategies that future research interventions and subsequent program can employ.

The findings of this review underscore the critical role of culturally-tailored strategies in engaging racialized newcomer populations in health promotion interventions. Across a variety of settings, from Canada to the United States, Europe and Australia, the use of community-based strategies and culturally congruent health workers was shown to be essential for ensuring high recruitment, retention and adherence rates. These findings align with the growing recognition that health interventions must be more than just accessible; they must also be relevant to the cultural, social, and emotional contexts of the participants.

Community health workers play a key role in delivering health promotion programs through delivering “culturally-appropriate health education, assist[ing] in translation or interpretation, and advocat[ing] for individual and community health needs, providing vital social support and capacity-building to ensure sustainable health improvements in their communities” [33, p. 206]. Involving community health workers/staff of the same racialized group is an effective strategy for delivering health promotion interventions to racialized groups.

Further, social networks, whether through peer support groups or community partnerships, were found to be invaluable for sustaining participation over time. The use of social media and virtual check-ins, as implemented during the COVID-19 pandemic in studies like Wieland et al. [68], also emerged as a way to maintain engagement when traditional in-person formats were not possible. These findings highlight the need to adapt engagement strategies to changing circumstances while ensuring that the sense of community and support remains intact.

Integrating culturally relevant references and activities into interventions were also key to ensuring programming met the needs of participants and fostered a recognizable environment in which to participate. Ensuring the newcomer community is involved throughout the entire process by using community-based participatory research principles emerged as a particularly promising approach, and researchers and policy makers should take care to engage in community-based strategies wherever possible.

Acknowledging barriers to participation and actively developing strategies to mitigate such challenges can prove helpful to effectively engage newcomers in CDPM programming. Incorporating incentives into programs, such as vouchers to offset costs and recognize participants’ time have been helpful, as have developing flexible delivery frameworks, incorporating technology to support participation, and developing materials and supports that ensure language is not a barrier to engagement. As future CDPM programs for newcomers are developed, care should be taken to ensure that barriers newcomers face to participating in such programs are adequately assessed and accounted for.

### Strengths and Limitations

This scoping review followed a rigorous framework [17, 18], ensuring systematic study selection. A comprehensive search strategy, developed in collaboration with an academic librarian, included multiple databases (Embase, MEDLINE, CINAHL, Scopus), maximizing study capture. Independent reviewers at each stage enhanced reliability, and iterative team discussions minimized biases. The use of the Elicit AI tool for data extraction streamlined the process, and pilot testing ensured accuracy. Verification of this extraction was also completed with pairs of reviewers, further enhancing rigor. As use of AI tools in research continues to be developed, this study offers an innovative approach to facilitate labour-intensive research processes. Descriptive analysis provided a clear overview of engagement strategies and intervention trends. However, this review’s language restriction to English and exclusion of non-empirical studies may have missed relevant studies for inclusion. The exclusion of migrants, temporary foreign workers, and international students as well as small sample sizes in some studies also limits the generalizability of findings. Additionally, the descriptive synthesis does not allow for deep statistical analysis, preventing conclusions on the effectiveness of specific interventions. This was outside the scope of the present review and should be considered in future research.

## Conclusion

The findings of this review underscore the multifaceted strategies required to effectively engage racialized newcomer populations in health promotion interventions. The key to success lies not only in the cultural relevance of the interventions but also in community involvement, trust-building, and the removal of as many barriers as possible. Community health workers, particularly those from the same racialized group, remain central to achieving these outcomes. However, addressing structural issues such as accessibility, economic support, and technological barriers are crucial to achieving equitable health outcomes for equity-deserving, racialized newcomer populations. Future research should continue to explore how to optimize these strategies while also ensuring their scalability and sustainability across diverse community contexts.

## Data Availability

All data produced in the present study are available upon reasonable request to the authors.

## Acknowledgements

Meagan Stanley, MLIS, Academic Librarian at Western University

## Statements and Declarations

### Competing Interests

This study was funded by Public Health Agency of Canada (PHAC) T2D Prevention Challenge.

### Ethics Approval

Ethics approval was not required for this review.

### Author Contributions

Conceptualization: Marisa L. Kfrerer, Dr. Robert J Petrella, Dr. Dawn P Gill; Literature Search: Megan Stanley; Data Analysis: Marisa L. Kfrerer, Ariana Tang, Paul Aspinall, Justin Ovadia; Writing: Marisa L. Kfrerer, Dr. Robert J Petrella, Dr. Dawn P Gill

## Appendix 1

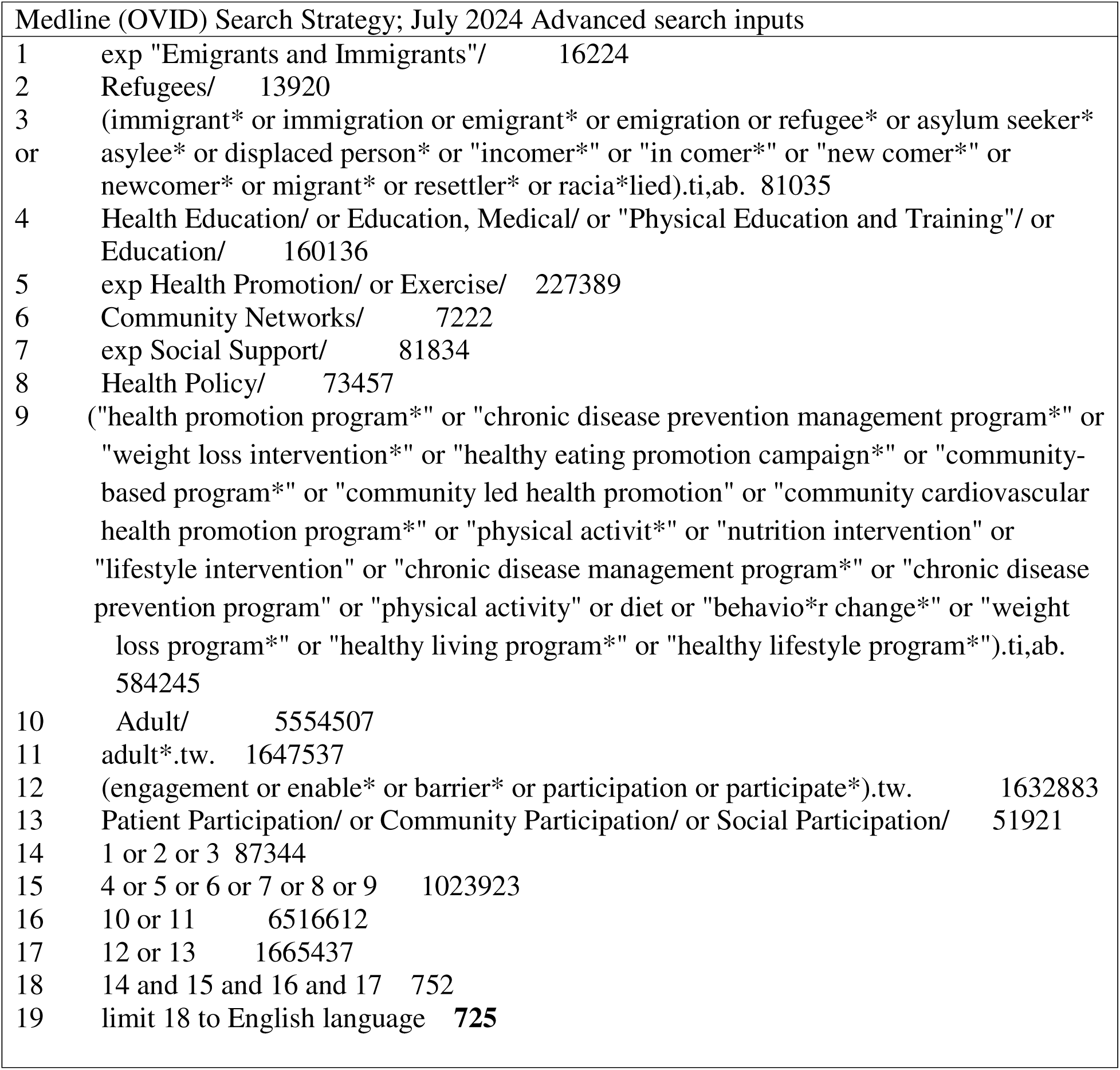

| Medline (OVID) Search Strategy; July 2024 Advanced search inputs |  |  |
| --- | --- | --- |
| 1 | exp "Emigrants and Immigrants"/ | 16224 |
| 2 | Refugees/ | 13920 |
| 3 | (immigrant* or immigration or emigrant* or emigration or refugee* or asylum seeker* or asylee* or displaced person* or "incomer*" or "in comer*" or "new comer*" or newcomer* or migrant* or resettler* or racia*lied).ti,ab. | 81035 |
| 4 | Health Education/ or Education, Medical/ or "Physical Education and Training"/ or Education/ | 160136 |
| 5 | exp Health Promotion/ or Exercise/ | 227389 |
| 6 | Community Networks/ | 7222 |
| 7 | exp Social Support/ | 81834 |
| 8 | Health Policy/ | 73457 |
| 9 | ("health promotion program*" or "chronic disease prevention management program*" or "weight loss intervention*" or "healthy eating promotion campaign*" or "community-based program*" or "community led health promotion" or "community cardiovascular health promotion program*" or "physical activit*" or "nutrition intervention" or "lifestyle intervention" or "chronic disease management program*" or "chronic disease prevention program" or "physical activity" or diet or "behavio*r change*" or "weight loss program*" or "healthy living program*" or "healthy lifestyle program*").ti,ab. | 584245 |
| 10 | Adult/ | 5554507 |
| 11 | adult*.tw. | 1647537 |
| 12 | (engagement or enable* or barrier* or participation or participate*).tw. | 1632883 |
| 13 | Patient Participation/ or Community Participation/ or Social Participation/ | 51921 |
| 14 | 1 or 2 or 3 | 87344 |
| 15 | 4 or 5 or 6 or 7 or 8 or 9 | 1023923 |
| 16 | 10 or 11 | 6516612 |
| 17 | 12 or 13 | 1665437 |
| 18 | 14 and 15 and 16 and 17 | 752 |
| 19 | limit 18 to English language | **725** |

## Notes

### Competing Interest Statement

The authors have declared no competing interest.

